# High-Throughput Safety Signal Detection for Sodium-Glucose Cotransporter-2 Inhibitors in Type 2 Diabetes

**DOI:** 10.1101/2025.11.25.25341015

**Authors:** Hao Dai, Yao An Lee, Jiang Bian, Jingchuan Guo

## Abstract

**Objective:** To systematically identify potential safety signals associated with sodium-glucose cotransporter-2 inhibitors (SGLT2i) in patients with type 2 diabetes (T2D) using a high-throughput target trial emulation pipeline applied to real-world data.

**Methods:** We utilized electronic health record (EHR) data from the OneFlorida+ Data Trust (2012–2023). Using a retrospective new-user cohort design, we compared SGLT2i initiators to initiators of other second-line glucose-lowering drugs (GLDs). We evaluated risk across all ICD-10-CM 4-digit diagnosis codes. A semi-Bayesian shrinkage method was employed to adjust hazard ratios (HR) and p-values to account for multiple testing and stabilize estimates for rare outcomes.

**Results:** The analysis identified several statistically significant safety signals. Validating our method, we observed known adverse events such as candidiasis of the vulva and vagina (HR=1.64) and other urogenital fungal infections. We also detected potential signals for conditions such as viral conjunctivitis and melanocytic nevi.

**Conclusion:** This high-throughput screening effectively identified both known and potential new safety signals for SGLT2i. The use of semi-Bayesian shrinkage provides a robust framework for post-marketing surveillance in large healthcare databases.

## Introduction

Sodium-glucose cotransporter-2 inhibitors (SGLT2i) have become a cornerstone in the management of type 2 diabetes (T2D), offering distinct benefits in glycemic control, weight loss, and cardiovascular and renal protection.^1^ Despite their widespread use and demonstrated efficacy in randomized controlled trials (RCTs), the full safety profile of SGLT2i in diverse, real-world populations remains an area of active investigation. RCTs are often limited by strict inclusion criteria, relatively small sample sizes, and short follow-up durations, which may preclude the detection of rare or long-term adverse events.

Post-marketing surveillance is critical to bridge this gap. Traditional pharmacovigilance relies heavily on spontaneous reporting systems, which are prone to underreporting and lack denominator data. Conversely, hypothesis-free “high-throughput” screening using large-scale electronic health records (EHR) allows for the systematic evaluation of thousands of potential adverse outcomes simultaneously.^2^ However, such massive multiple testing increases the risk of false-positive findings.

To address this challenge, we designed a high-throughput target trial emulation pipeline. We applied this pipeline to the OneFlorida+ Data Trust, a large, diverse repository of longitudinal EHR data. To rigorously control for false discoveries while maintaining sensitivity for rare events, we incorporated semi-Bayesian shrinkage estimation.^3^ This study aims to systematically scan for and quantify safety signals associated with SGLT2i use compared to other second-line glucose-lowering drugs (GLDs) in a real-world T2D cohort.

## Methods

### Data source

Data were drawn from the centralized repository maintained by the OneFlorida+ Clinical Research Consortium. OneFlorida+ links longitudinal EHRs with ancillary sources and includes demographics, diagnoses, medications, procedures, vital signs, and laboratories across participating systems. Source data covered January 1, 2012–June 30, 2023; cohort spanned January 1, 2016–June 30, 2023.

### Study design

This study employed a retrospective cohort design with a new-user approach. An intention-to-treat analysis was conducted to compare sodium–glucose cotransporter-2 inhibitors (SGLT2is) with other second-line GLDs, in terms of sulfonylureas, thiazolidinediones, dipeptidyl peptidase-4 inhibitors (DPP-4i), α-glucosidase inhibitors, and meglitinides, on the risk of diagnosis-specific outcomes defined at the ICD-10-CM 4-digit level among adults with T2D. A uniform time-to-event specification, including common covariate adjustment and censoring rules—was applied across all endpoints to enable high-throughput estimation. Exposure definitions are summarized in **Table 1**, and the overall design is illustrated in **Figure 1**. The study was approved by the University of Florida Institutional Review Board (IRB202201196).

**Figure 1.**
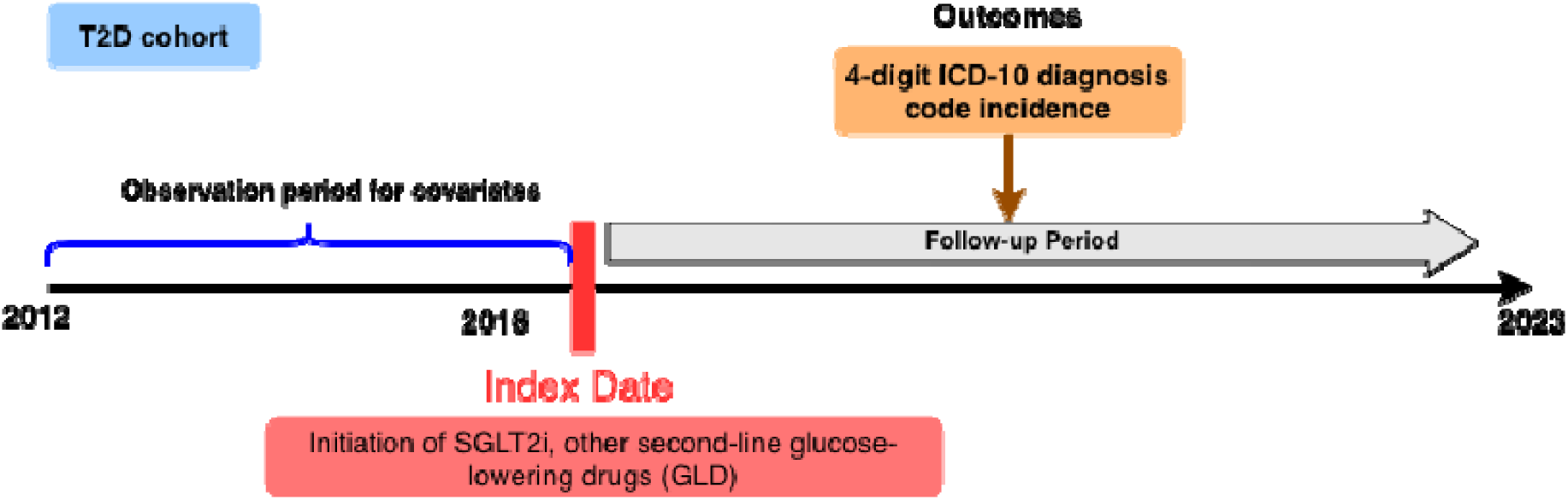
Overview of study design: new users of a SGLT2i, or other second-line glucose-lowering drug (GLD). Other second-line GLDs include sulfonylureas, thiazolidinediones, DPP4i, α-glucosidase inhibitors, or meglitinides

**Table 1.** Drugs of interest.

|  |  |  |
| --- | --- | --- |
| Drugs of interests | SGLT2is | Canagliflozin; Dapagliflozin; Empagliflozin; Ertugliflozin |
| Other second-line GLDs | DPP4is | Sitagliptin; Saxagliptin; Alogliptin; Linagliptin |
|  | Sulfonylureas | Glimepiride; Glyburide; Chlorpropamide; Glipizide; Tolbutamide; Tolazamide |
|  | Thiazolidinediones | Pioglitazone; Rosiglitazone |
|  | Meglitinides | Repaglinide; Nateglinide |
| | α-glucosidase inhibitors | Miglitol; Acarbose |

DPP4is, dipeptidyl peptidase 4 inhibitors; GLP-1RAs, glucagon-like peptide-1 receptor agonists; SGLT2is, sodium-glucose cotransporter-2 inhibitors; GLDs, glucose-lowering drugs.

### Eligibility criteria

We included people who initiated treatment with a SGLT2i or other second-line GLDs in OneFlorida+ between January 1, 2016 and June 30, 2023. Other second-line GLDs were selected as the comparator group to help reduce confounding by indication (**Table 1**).^4^ The index date was the first recorded prescription for an SGLT2 inhibitor or an active-comparator GLD without a previous prescription for either drug in the previous year. We excluded individuals who were younger than 18 years at index, had a diagnosis of type 1 diabetes, gestational diabetes, or end-stage renal disease (ESRD) on or before index, or had no clinical encounters prior to index. Operational definitions and code lists are provided in **Table 2**.

**Table 2.** Codes used for key variables.

| Outcome/key variables | ICD-9 | ICD-10 |
| --- | --- | --- |
| Type 2 diabetes | 250.x0; 250.x2 | E11 |
| Type 1 diabetes | 250.x1; 250.x3 | E10 |
| Gestational diabetes | 648.0x | O24 |
| End-Stage Renal Disease | 585.6 | N18.6 |

### Outcome and follow-up

The outcome set comprised incident ICD-10-CM 4-digit diagnosis codes recorded during follow-up. For each 4-digit code, the first post-index occurrence was considered the incident event; persons with that code present at baseline were excluded from that outcome’s risk set. Patients’ follow-up began at the index date and continued until the earliest occurrence of one of the following censoring events: the outcome of interest, death, loss to follow-up (the date of the last recorded clinical encounter), or the end of the study period (June 30, 2023).

### Baseline covariates

Baseline covariates encompassed a range of demographic, clinical, and pharmacological characteristics, as detailed in **Table 3**. Demographic variables included age, sex, race/ethnicity, smoking status, and insurance type. Clinical covariates comprised comorbid conditions (e.g., cardiovascular disease, cerebrovascular disease and neuropathy), clinical observations (e.g., BMI, blood pressure), and medication (e.g., insulin, opioids, and statins).

**Table 3.**
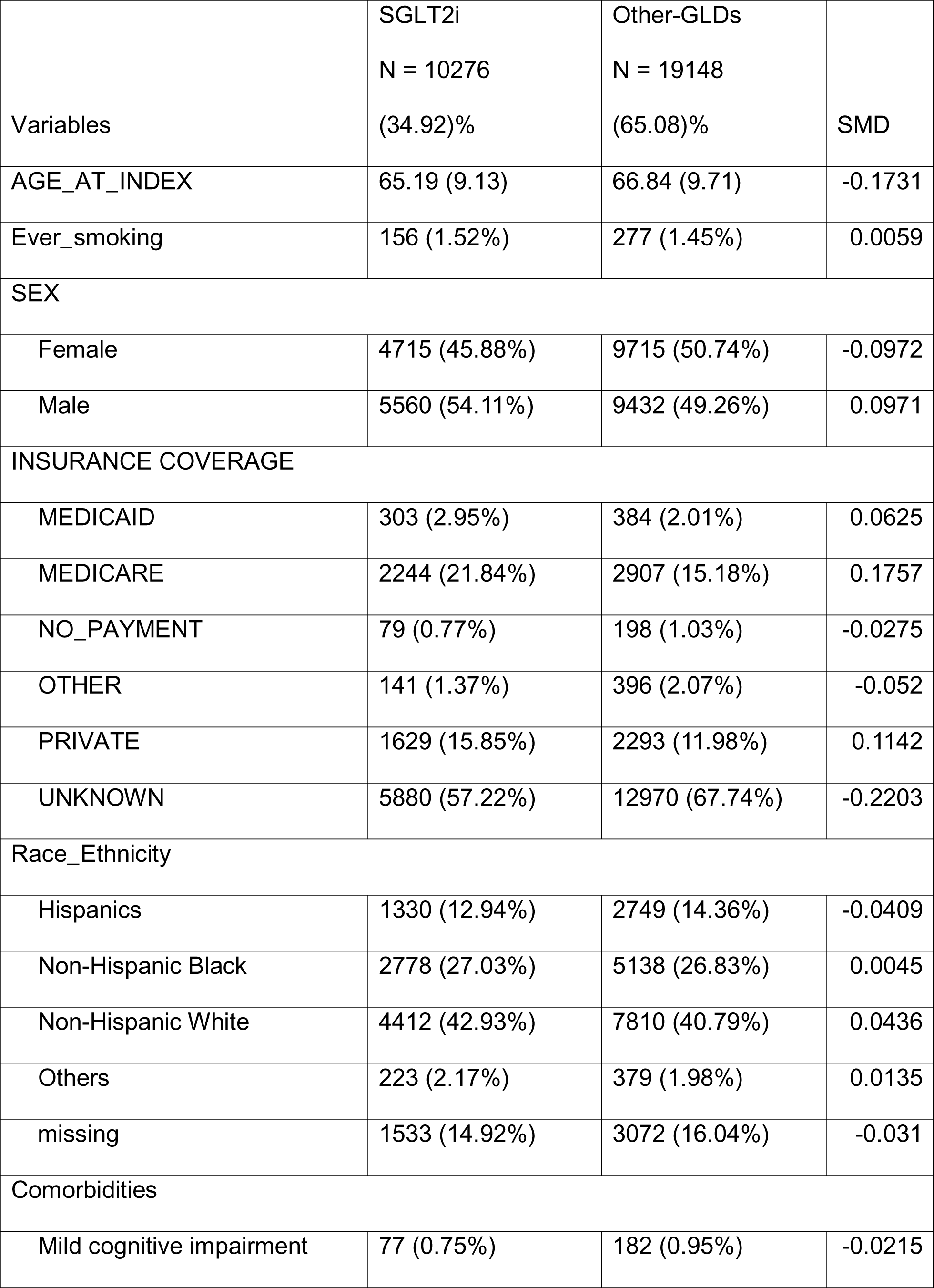
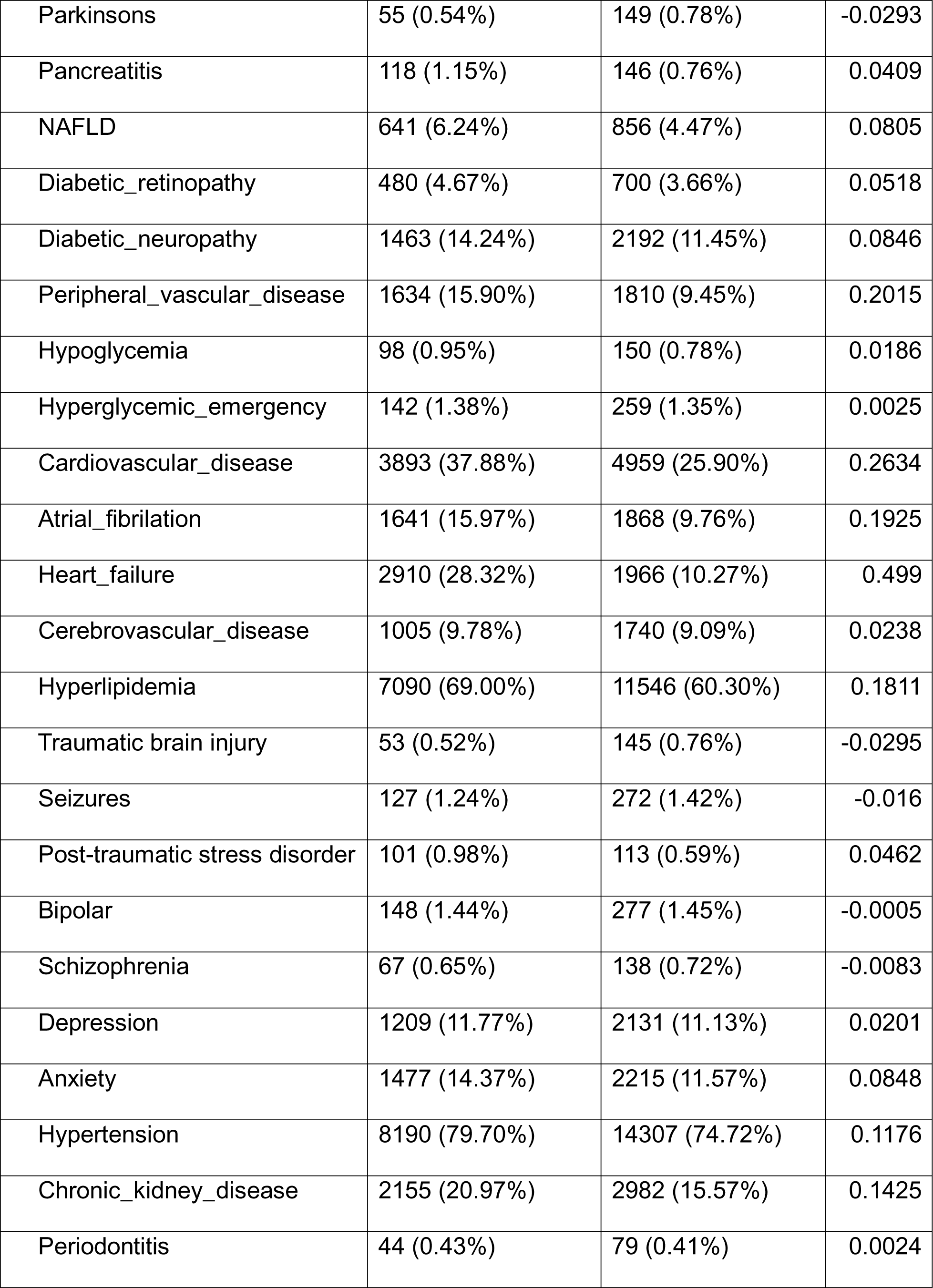
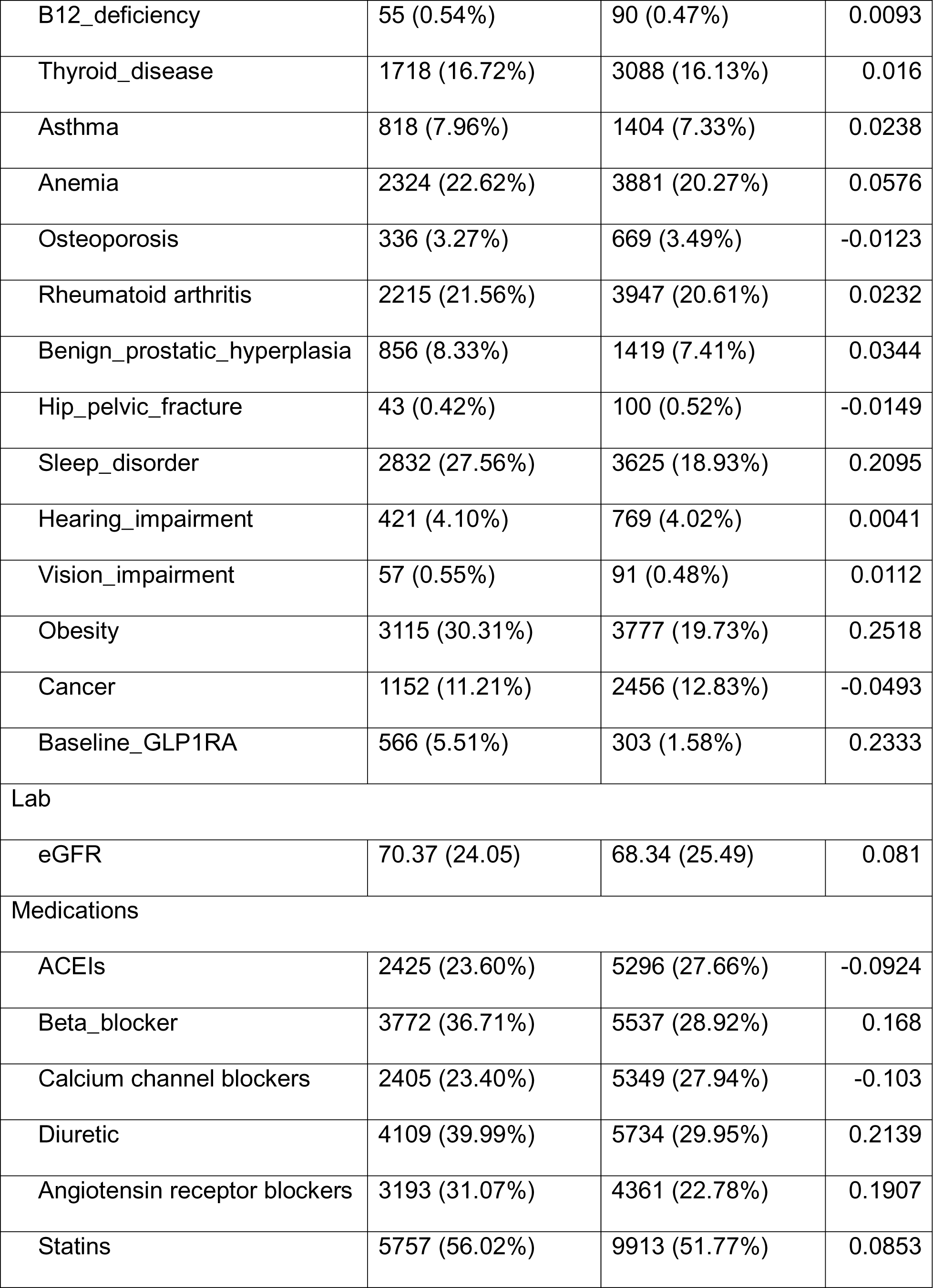
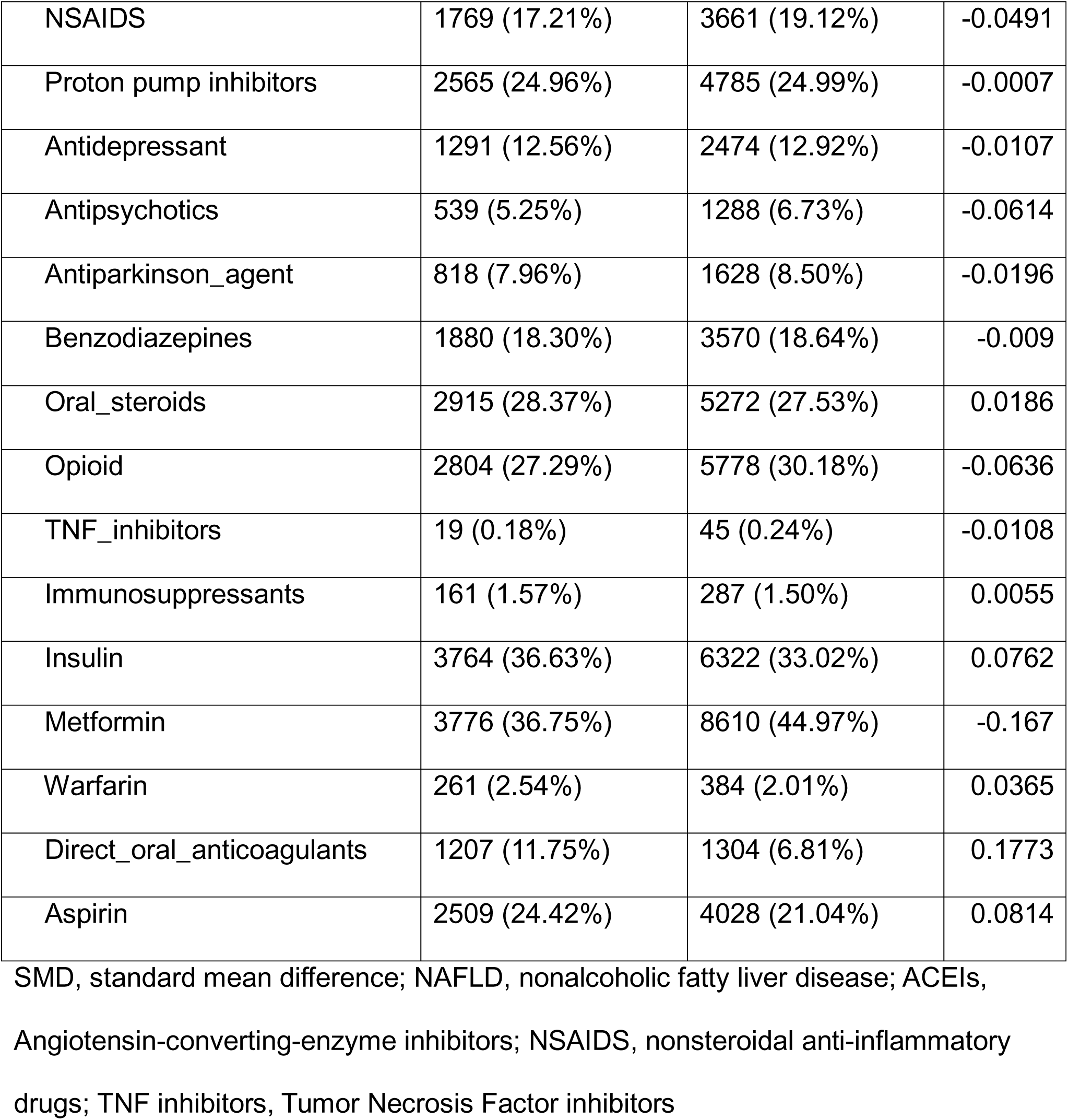
Baseline variable characteristics.

| Variables | SGLT2i<br>N = 10276<br>(34.92)% | Other-GLDs<br>N = 19148<br>(65.08)% | SMD |
| --- | --- | --- | --- |
| AGE_AT_INDEX | 65.19 (9.13) | 66.84 (9.71) | -0.1731 |
| Ever_smoking | 156 (1.52%) | 277 (1.45%) | 0.0059 |
| SEX |  |  |  |
| Female | 4715 (45.88%) | 9715 (50.74%) | -0.0972 |
| Male | 5560 (54.11%) | 9432 (49.26%) | 0.0971 |
| INSURANCE COVERAGE |  |  |  |
| MEDICAID | 303 (2.95%) | 384 (2.01%) | 0.0625 |
| MEDICARE | 2244 (21.84%) | 2907 (15.18%) | 0.1757 |
| NO_PAYMENT | 79 (0.77%) | 198 (1.03%) | -0.0275 |
| OTHER | 141 (1.37%) | 396 (2.07%) | -0.052 |
| PRIVATE | 1629 (15.85%) | 2293 (11.98%) | 0.1142 |
| UNKNOWN | 5880 (57.22%) | 12970 (67.74%) | -0.2203 |
| Race_Ethnicity |  |  |  |
| Hispanics | 1330 (12.94%) | 2749 (14.36%) | -0.0409 |
| Non-Hispanic Black | 2778 (27.03%) | 5138 (26.83%) | 0.0045 |
| Non-Hispanic White | 4412 (42.93%) | 7810 (40.79%) | 0.0436 |
| Others | 223 (2.17%) | 379 (1.98%) | 0.0135 |
| missing | 1533 (14.92%) | 3072 (16.04%) | -0.031 |
| Comorbidities |  |  |  |
| Mild cognitive impairment | 77 (0.75%) | 182 (0.95%) | -0.0215 |
| Parkinsons | 55 (0.54%) | 149 (0.78%) | -0.0293 |
| Pancreatitis | 118 (1.15%) | 146 (0.76%) | 0.0409 |
| NAFLD | 641 (6.24%) | 856 (4.47%) | 0.0805 |
| Diabetic_retinopathy | 480 (4.67%) | 700 (3.66%) | 0.0518 |
| Diabetic_neuropathy | 1463 (14.24%) | 2192 (11.45%) | 0.0846 |
| Peripheral_vascular_disease | 1634 (15.90%) | 1810 (9.45%) | 0.2015 |
| Hypoglycemia | 98 (0.95%) | 150 (0.78%) | 0.0186 |
| Hyperglycemic_emergency | 142 (1.38%) | 259 (1.35%) | 0.0025 |
| Cardiovascular_disease | 3893 (37.88%) | 4959 (25.90%) | 0.2634 |
| Atrial_fibrillation | 1641 (15.97%) | 1868 (9.76%) | 0.1925 |
| Heart_failure | 2910 (28.32%) | 1966 (10.27%) | 0.499 |
| Cerebrovascular_disease | 1005 (9.78%) | 1740 (9.09%) | 0.0238 |
| Hyperlipidemia | 7090 (69.00%) | 11546 (60.30%) | 0.1811 |
| Traumatic brain injury | 53 (0.52%) | 145 (0.76%) | -0.0295 |
| Seizures | 127 (1.24%) | 272 (1.42%) | -0.016 |
| Post-traumatic stress disorder | 101 (0.98%) | 113 (0.59%) | 0.0462 |
| Bipolar | 148 (1.44%) | 277 (1.45%) | -0.0005 |
| Schizophrenia | 67 (0.65%) | 138 (0.72%) | -0.0083 |
| Depression | 1209 (11.77%) | 2131 (11.13%) | 0.0201 |
| Anxiety | 1477 (14.37%) | 2215 (11.57%) | 0.0848 |
| Hypertension | 8190 (79.70%) | 14307 (74.72%) | 0.1176 |
| Chronic_kidney_disease | 2155 (20.97%) | 2982 (15.57%) | 0.1425 |
| Periodontitis | 44 (0.43%) | 79 (0.41%) | 0.0024 |
| B12_deficiency | 55 (0.54%) | 90 (0.47%) | 0.0093 |
| Thyroid_disease | 1718 (16.72%) | 3088 (16.13%) | 0.016 |
| Asthma | 818 (7.96%) | 1404 (7.33%) | 0.0238 |
| Anemia | 2324 (22.62%) | 3881 (20.27%) | 0.0576 |
| Osteoporosis | 336 (3.27%) | 669 (3.49%) | -0.0123 |
| Rheumatoid arthritis | 2215 (21.56%) | 3947 (20.61%) | 0.0232 |
| Benign_prostatic_hyperplasia | 856 (8.33%) | 1419 (7.41%) | 0.0344 |
| Hip_pelvic_fracture | 43 (0.42%) | 100 (0.52%) | -0.0149 |
| Sleep_disorder | 2832 (27.56%) | 3625 (18.93%) | 0.2095 |
| Hearing_impairment | 421 (4.10%) | 769 (4.02%) | 0.0041 |
| Vision_impairment | 57 (0.55%) | 91 (0.48%) | 0.0112 |
| Obesity | 3115 (30.31%) | 3777 (19.73%) | 0.2518 |
| Cancer | 1152 (11.21%) | 2456 (12.83%) | -0.0493 |
| Baseline_GLP1RA | 566 (5.51%) | 303 (1.58%) | 0.2333 |
| Lab |  |  |  |
| eGFR | 70.37 (24.05) | 68.34 (25.49) | 0.081 |
| Medications |  |  |  |
| ACEIs | 2425 (23.60%) | 5296 (27.66%) | -0.0924 |
| Beta_blocker | 3772 (36.71%) | 5537 (28.92%) | 0.168 |
| Calcium channel blockers | 2405 (23.40%) | 5349 (27.94%) | -0.103 |
| Diuretic | 4109 (39.99%) | 5734 (29.95%) | 0.2139 |
| Angiotensin receptor blockers | 3193 (31.07%) | 4361 (22.78%) | 0.1907 |
| Statins | 5757 (56.02%) | 9913 (51.77%) | 0.0853 |
| NSAIDS | 1769 (17.21%) | 3661 (19.12%) | -0.0491 |
| Proton pump inhibitors | 2565 (24.96%) | 4785 (24.99%) | -0.0007 |
| Antidepressant | 1291 (12.56%) | 2474 (12.92%) | -0.0107 |
| Antipsychotics | 539 (5.25%) | 1288 (6.73%) | -0.0614 |
| Antiparkinson_agent | 818 (7.96%) | 1628 (8.50%) | -0.0196 |
| Benzodiazepines | 1880 (18.30%) | 3570 (18.64%) | -0.009 |
| Oral_steroids | 2915 (28.37%) | 5272 (27.53%) | 0.0186 |
| Opioid | 2804 (27.29%) | 5778 (30.18%) | -0.0636 |
| TNF_inhibitors | 19 (0.18%) | 45 (0.24%) | -0.0108 |
| Immunosuppressants | 161 (1.57%) | 287 (1.50%) | 0.0055 |
| Insulin | 3764 (36.63%) | 6322 (33.02%) | 0.0762 |
| Metformin | 3776 (36.75%) | 8610 (44.97%) | -0.167 |
| Warfarin | 261 (2.54%) | 384 (2.01%) | 0.0365 |
| Direct_oral_anticoagulants | 1207 (11.75%) | 1304 (6.81%) | 0.1773 |
| Aspirin | 2509 (24.42%) | 4028 (21.04%) | 0.0814 |

SMD, standard mean difference; NAFLD, nonalcoholic fatty liver disease; ACEIs, Angiotensin-converting-enzyme inhibitors; NSAIDS, nonsteroidal anti-inflammatory drugs; TNF inhibitors, Tumor Necrosis Factor inhibitors

### High-throughput modeling and semi-Bayesian shrinkage

Building on the previously defined cohort and exposure framework, we implemented a high-throughput outcome-wide screening pipeline across ICD-10-CM 4-digit diagnoses, treating each code as an independent endpoint within a unified modeling and semi-Bayesian shrinkage framework. We estimated a propensity score for treatment (SGLT2i initiation vs alternative second-line GLDs) via logistic regression on the prespecified covariate list, constructed stabilized inverse-probability weights, and trimmed weights at the 0.5th and 99.5th percentiles to mitigate extreme influence.^5^ We then computed weighted standardized mean differences (SMDs) for each covariate; any variable with an absolute weighted SMD > 0.20 was then included in the outcome-specific Cox model as an adjuster to account for residual imbalance.^56^ For each outcome we fit an IPTW-weighted Cox proportional hazards model with treatment as exposure and adjustment for any residual imbalance covariates; outcomes with zero events or non-convergent fits were skipped, and rows with missing model inputs were dropped. To stabilize the many effect estimates derived across thousands of outcomes and reduce the risk of false positives (as recognized in prior large-scale screening frameworks such as the OHDSI high-throughput paradigm), we applied semi-Bayesian shrinkage to the vector of treatment log-hazard estimates.^7^ Specifically, for outcome *j* with estimate 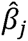 and standard error *se_j_*, we assumed 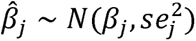 and *β_j_ ∼ N*(*µ, ꚍ*^2^), where *ꚍ*^2^ is a fixed prior variance (default 0.25) and *µ* is either zero (“zero-target”) or the inverse-variance weighted global mean of the raw estimates (“global-target”). The posterior mean and variance were calculated analytically and converted into shrinkage-adjusted hazard ratios, with corresponding Wald 95% confidence intervals. We also reported the shrinkage factor (*ꚍ*^2^/(*ꚍ*^2^ + *se_j_*^2^)), semi-Bayes p-values based on the posterior, and applied Benjamini–Hochberg false discovery rate (FDR) correction across outcomes. Outcomes were flagged as signals when the lower bound of the shrinkage-adjusted 95% interval exceeded 1.00. Sensitivity analyses varied *ꚍ*^2^ and the choice of *µ* (zero vs global) and, for a subset of endpoints, we confirmed results with a Bayesian sampling approach implemented in PyMC. All intermediate artifacts—propensity scores, weights, diagnostics, model convergence flags and summary tables—were stored to ensure full reproducibility and auditability of the high-throughput pipeline.

### Collinearity analysis

To assess redundancy among the flagged 4-digit outcomes, we quantified pairwise association using the Phi coefficient (φ) for binary outcomes. For each outcome pair, we constructed a 2x 2 contingency table among individuals free of both outcomes at baseline and computed

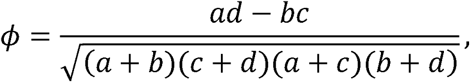

where *a*–*d* are joint cell counts. When a strict 2×2 table was not feasible (e.g., zero cells), we used Cramér’s V as a generalized measure. A custom Python workflow iteratively computed φ across all pairs to assemble a symmetric Φ-matrix. High collinearity was defined as | *ø* |≥ 0.70; pairs with | ø |> 0.85 were considered *extremely* correlated. For high-correlation pairs, we summarized prevalence, overlap (both/only/neither), and risk-set size, generated a heatmap for visual QC (upper triangle masked; high-correlation cells outlined), and then retained one representative code per cluster, prioritizing the most specific 4-digit diagnosis with clear clinical interpretability.

## Results

### Cohort Selection and Baseline Characteristics

A total of 29,425 patients with type 2 diabetes who met the eligibility criteria were included in the study cohort. Among them, 10,277 patients initiated treatment with SGLT2 inhibitors (SGLT2i), and 19,148 patients initiated other second-line glucose-lowering drugs (GLDs).

Before propensity score weighting, some imbalances were observed between the two study groups (**Table 1**). Compared to the comparator group, SGLT2i initiators were generally younger (mean age: 58.7 vs. 61.3 years), had a higher body mass index (BMI) (mean: 33.8 vs. 31.4 kg/m²), and had a higher baseline HbA1c (mean: 8.35% vs. 7.92%). The prevalence of certain comorbidities, such as hyperlipidemia (76.8% vs. 69.8%), was also higher in the SGLT2i group. In terms of concomitant medications, SGLT2i users were more likely to be treated with metformin (79.0% vs. 64.9%) and statins (56.0% vs. 51.8%).

After applying overlap weighting, all measured baseline characteristics were well-balanced between the SGLT2i and comparator groups, with standardized mean differences (SMD) for all covariates falling below 0.1, indicating successful adjustment for observed confounders.

### High-Throughput Safety Signal Detection

We performed a systematic, high-throughput screening across all available ICD-10-CM 4-digit diagnosis codes to identify potential safety signals. **Figure 2** illustrates the distribution of the estimated HR across different organ system categories.

**Figure 2.**
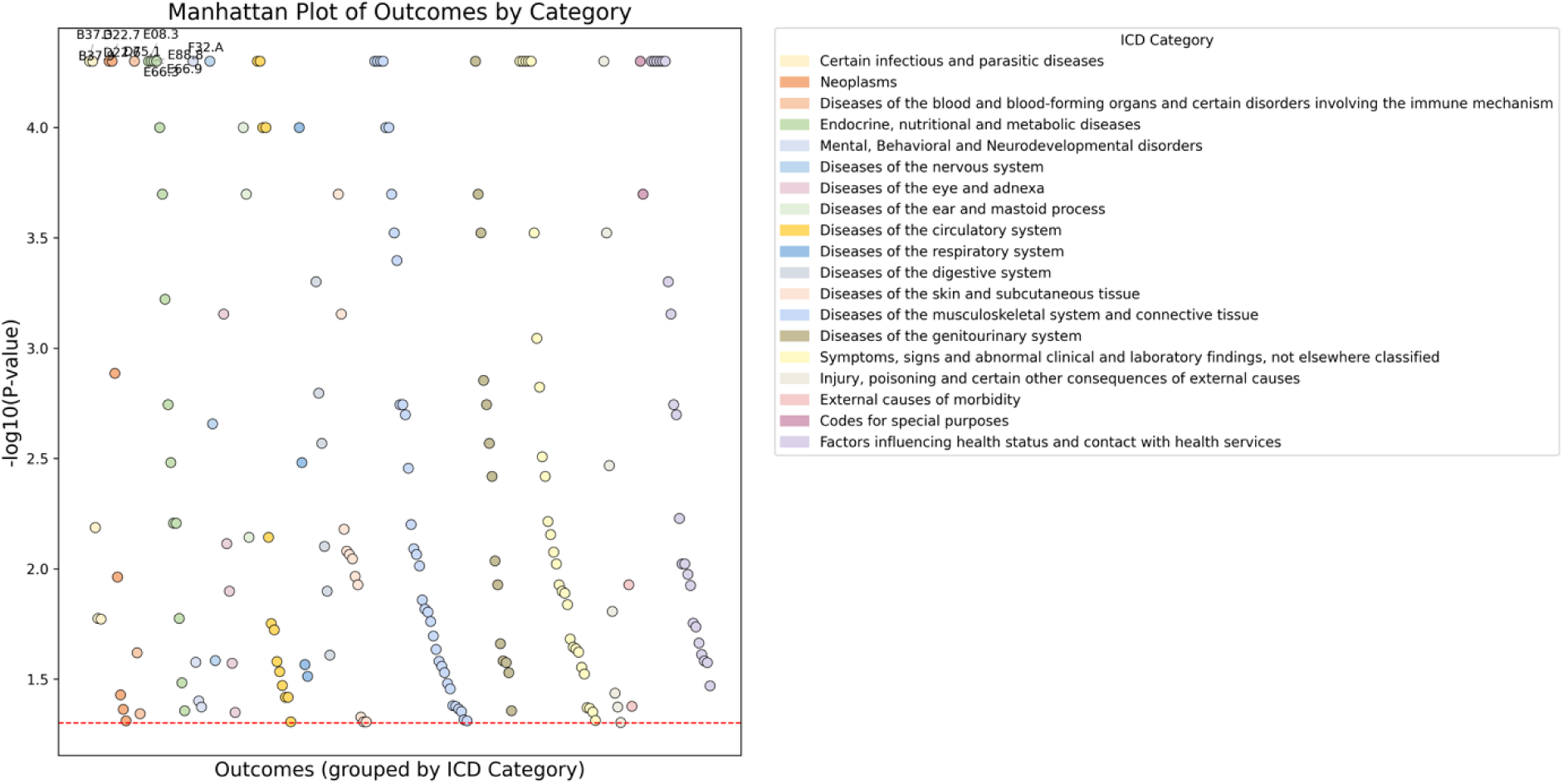

### Identification of Specific Safety Signals

Using semi-Bayesian shrinkage to penalize unstable estimates and control for multiple testing, we identified several diagnosis codes with a shrinkage-adjusted HR significantly greater than 1. Consistent with well-established clinical trial data, our pipeline successfully identified genital mycotic infections as the most prominent safety signal, validated by high E-values which suggest that unmeasured confounding is unlikely to explain these associations. Specifically, SGLT2i use was associated with elevated risks of candidiasis of the vulva and vagina (HR 1.64; 95% CI 1.42–1.90), unspecified candidiasis (HR 1.92; 95% CI 1.63–2.26), and candidiasis of other urogenital sites (HR 1.35; 95% CI 1.06–1.73). Beyond these known genitourinary infections, the high-throughput analysis highlighted several other statistically significant associations that may warrant further investigation, including viral conjunctivitis (HR 1.58; 95% CI 1.09–2.31), other specified bacterial intestinal infections (HR 1.40; 95% CI 1.10–1.78), and melanocytic nevi of the trunk (HR 1.34; 95% CI 1.12–1.60). Furthermore, signals related to health service utilization were detected within the “Factors influencing health status” category—such as screening for malignant neoplasm of respiratory organs (HR 1.53; 95% CI 1.29–1.81), presence of endocrine implants (HR 1.62; 95% CI 1.19–2.19), and screening for blood/immune disorders (HR 1.35; 95% CI 1.14–1.60)—which likely reflect surveillance bias or increased healthcare contact frequency rather than direct pharmacological toxicity.

### Robustness of Estimates

The semi-Bayesian shrinkage method was applied to all estimates. For the primary signals (e.g., Candidiasis), the shrinkage factor was high (>0.89), indicating that the raw data provided strong evidence with stable variance. For rarer outcomes (e.g., Viral conjunctivitis), the shrinkage factor was lower (∼0.41), demonstrating the method’s conservative approach to effectively reduce false positives in low-prevalence outcomes.

## Discussion

In this study, we successfully implemented a high-throughput target trial emulation pipeline to screen for safety signals associated with SGLT2i use in a large, real-world population. By employing semi-Bayesian shrinkage, we identified significant associations that align with known adverse drug reactions while highlighting potential areas for future research.

### Interpretation of Findings

The detection of genital mycotic infections (candidiasis) validates the sensitivity of our pipeline. SGLT2 inhibitors induce glycosuria, creating a favorable environment for fungal growth in the genitourinary tract. This is the most well-documented side effect of the class, reported consistently across RCTs and observational studies.^8^ The magnitude of the risk observed in our study (HR ∼1.6–1.9) is consistent with the literature, confirming that our methods effectively replicate known causal associations.

We also observed a signal for **viral conjunctivitis** and **melanocytic nevi**. While less commonly associated with SGLT2i, skin and subcutaneous tissue disorders have been reported in pharmacovigilance databases.^9^ The signal for nevi could potentially be attributed to surveillance bias (patients on newer therapies may receive more frequent physical exams) or chance, highlighting the need for specific validation studies. Similarly, signals related to specific bacterial infections (e.g., intestinal) may reflect the known risk of volume depletion or immunomodulation, though confounding by indication remains a possibility.

### Strengths and Limitations

A major strength of this study is the use of the OneFlorida+ database, which provides a diverse and representative sample. The use of semi-Bayesian shrinkage is a methodological strength, allowing us to screen thousands of outcomes without generating an excessive number of false positives.^3^ Limitations include the observational nature of the data, which subjects the analysis to residual confounding despite the new-user design. Additionally, ICD-10 coding may lack the precision of adjudicated outcomes used in clinical trials.

### Conclusion

Our high-throughput pipeline confirmed the known risk of conditions associated with SGLT2i and generated hypotheses regarding other potential adverse events. This approach demonstrates the utility of applying advanced statistical methods to real-world data for comprehensive post-marketing drug safety surveillance.

## Author Contributions

Dr. J. Guo is the guarantor of this work and, as such, had full access to all the data in the study and takes responsibility for the integrity of the data and the accuracy of the data analysis.

Concept and design: J. Guo, and J. Bian.

Acquisition, analysis, or interpretation of data: All authors.

Drafting of the manuscript: H. Dai, Y. Lee

Critical review of the manuscript for important intellectual content: All authors.

Statistical analysis: H. Dai and Y. Lee

Obtained funding: J. Guo.

Supervision: J. Guo and J. Bian.

## Data Availability Statement

Data set Available through OneFlorida+ Clinical Research Network (email)

